# RCC-AID: Renal Cell Carcinoma AI Dataset for Medical Imaging Research

**DOI:** 10.64898/2026.04.22.26351451

**Authors:** Sarah de Boer, Hartmut Häntze, Sebastian Ziegelmayer, Tommaso Russo, Bram van Ginneken, Mathias Prokop, Keno Bressem, Alessa Hering

## Abstract

Contrast-enhanced computed tomography (CT) is central to the diagnosis, staging, and follow-up of patients with renal cell carcinoma (RCC). As artificial intelligence research into computer-aided solutions continues to grow, the need for curated and annotated datasets becomes increasingly important. Imaging-based artificial intelligence studies often need lesion annotations that are not consistently available. The Cancer Genome Atlas (TCGA) datasets are widely used for model training and validation. However, access to public annotations of lesions is limited, which limits reproducibility and comparability of the published research. To address this gap, we screened 1,915 CT scans from three TCGA-RCC databases and, following a meta-data-based exclusion step, used an automated segmentation model to generate initial kidney and lesion masks. Next, we conducted a reader study with all papillary (n=56), chromophobe (n=27) and 200 randomly selected clear cell RCC cases. Two trained students performed quality checks, corrections, and additional annotation of tumors and cysts, with uncertain cases reviewed by a board-certified radiologist. After data exclusion and quality control, a final cohort of 129 annotated CT scans from 91 patients (24 female, 67 male; mean age 56 years) was retained, including 85 clear cell, 26 papillary and 18 chromophobe RCC cases. Images and voxel-level annotations of kidneys and lesions are openly available at https://zenodo.org/records/20719257. By open-sourcing these annotations, we aim to foster accessible, reproducible AI research in renal cell carcinoma. RCC-AID provides a reusable open resource dataset for segmentation, detection, subtype classification, radiomics, and multimodal RCC research.

## 1. Background

**Renal** cell carcinoma (RCC) is the most common form of kidney cancer, with clear cell (70%), papillary (10–15%), and chromophobe (5%) carcinoma the main histologic subtypes (Moch et al., 2022). Imaging is essential across the RCC care pathway, from diagnosis through treatment planning and decisions to post-treatment surveillance. Computed tomography (CT) is the primary modality used for RCC imaging, due to its wide availability, cost-effectiveness, and high image quality (Abou Elkassem et al., 2021; Sankineni et al., 2016; Rossi et al., 2018; Baytok et al., 2025).

The Cancer Genome Atlas (TCGA) is a widely used public resource for multimodal cancer research, providing matched genomic, histopathological, and clinical data, and, via linked repositories such as The Cancer Imaging Archive (TCIA), radiological imaging data. For RCC, TCGA includes three subtype-specific cohorts: TCGA-KIRC (clear cell) (Akin et al., 2016), TCGA-KICH (chromophobe) (Linehan et al., 2016b), and TCGA-KIRP (papillary) (Linehan et al., 2016a). These cohorts are commonly used for methodological development in machine learning and deep learning, as well as for retrospective clinical and translational analyses (e.g. Nguyen et al. (2018); Zhu et al. (2016); Doshi et al. (2021); Gao et al. (2024); Liu et al. (2023)).

The RCC cohorts in TCGA have enabled a broad range of computational imaging studies, spanning detection and subtype classification (Liu et al., 2023; Sitanaboina et al., 2023; Uhm et al., 2021), tumor staging, grading, and survival prediction (Lin et al., 2020; Wang et al., 2023; Moitra and Mandal, 2022; Yuan et al., 2025), radiomics and texture analysis (Doshi et al., 2021; Gao et al., 2024; Lin et al., 2021), and cross-modal image registration (Hering et al., 2023; Siebert et al., 2022). Annotation requirements vary by objective: detection and segmentation need voxel-level tumor delineations, staging/grading rely on case-level labels, radiomics needs defined regions of interest, and registration needs corresponding anatomical landmarks or voxel-level volume segmentations rather than tumor types. These annotation types are not interchangeable, yet are frequently built independently on the same images under different protocols.

Variability in imaging and annotation protocols affects reproducibility and comparability of studies using TCGA cohorts, particularly as annotation resources are often generated independently and are not always shared in sufficient detail for reuse. Consequently, studies based on the same TCGA cohort may rely on different reference standards, which can complicate cross-study comparison and limit opportunities for aggregation.

Although public annotations are essential for reproducible imaging AI research, voxel-level kidney and lesion annotations are not consistently available for the TCGA RCC imaging cohorts. Recent initiatives (Murugesan et al., 2024, 2025) have used AI-strategies to annotate certain TCGA cohorts, inclusing lung, prostate, kidney and breast. These efforts have expanded the availability of annotated cancer imaging data, but coverage remains incomplete for some RCC cohorts, including KICH and KIRP.

We address this gap by releasing RCC-AID, a publicly available dataset with voxel-level annotations of kidneys, primary solid tumors, and cysts in contrast-enhanced CT from three TCGA RCC cohorts, supplemented by the annotation of the contrast phase of the CT. By covering clear cell, papillary, and chromophobe RCC, RCC-AID is designed to support RCC-related tasks such as kidney and tumor segmentation, lesion detection, cyst identification, subtype classification, and radiomics analysis. By making this dataset publicly available, we aim to support more reproducible and accessible deep learning research in RCC.

## 2. Summary

RCC-AID is an open, annotated CT dataset designed to support reproducible AI research in renal cell carcinoma. We provide ready-to-use voxel-level annotations for three major RCC histologic subtypes: clear cell, papillary, and chromophobe, derived from the widely used TCGA cohorts. By consolidating and openly releasing these annotations, RCC-AID reduces redundant annotation efforts across studies, enables fair benchmarking, and fosters cross-study comparability.

Images are provided in NIfTI format (.nii.gz) alongside corresponding segmentation masks, suitable for direct use by standard deep learning pipelines such as nnU-Net (Isensee et al., 2021). Each scan includes a single contrast-enhanced CT volume with voxel-level labels for the kidney, primary solid tumor, and cysts where present. Accompanying metadata, including histologic subtype, patient sex, age, tumor size, and contrast phase of the CT, are provided as a structured CSV file.

No specialized infrastructure is required to use this dataset. Standard medical imaging libraries such as SimpleITK, ITK-SNAP, or 3D Slicer can be used to visualize and process the data.

## 3. Discussion

The renal cell carcinoma cohorts available through TCGA have been widely used in the literature for model development and validation. However, cross-study comparability is compromised when imaging data and annotated reference standards are not publicly available and different between studies. RCC-AID provides publicly available voxel-level annotations of kidneys, primary solid tumors, and cysts in contrast-enhanced CT scans from the TCGA-KIRC, TCGA-KIRP, and TCGA-KICH cohorts. In total, 129 CT scans from 91 patients are publicly released and immediately usable for AI model development and benchmarking.

Recent annotation efforts for TCGA RCC cohorts include the work Murugesan et al. (2024, 2025), which focused primarily on the KIRC clear cell cohort. RCC-AID complements these initiatives by extending coverage to all three major histologic subtypes, including the previously unannotated KICH and KIRP cohorts, and by providing annotations for both tumors and cysts.

Several limitations of this dataset should be taken into account when using it. The TCGA imaging data are heterogeneous in nature, with extensive variability in CT ac-RCC-AID: Renal Cell Carcinoma AI Dataset for Medical Imaging Research quisition parameters (e.g., slice thickness ranging from 0.6 to 5.0 mm). The annotations were initialized using an AI segmentation model and subsequently reviewed and corrected by trained student annotators, with radiologist review for uncertain cases. As formal inter-observer variability was not assessed, the masks should be regarded as quality-controlled research annotations rather than exhaustive expert-consensus annotations. For many downstream applications, lesion-level detection errors, such as false positives and missed tumors may be more relevant than small boundary differences in Dice similarity coefficient.

The dataset reflects known demographic and geographic biases inherent to the TCGA cohorts. The patient population is predominantly male (67 of 91 patients) and was recruited across North American academic medical centers, which may limit generalizability to other populations. Models trained on this cohort may underperform in populations differing in sex distribution, comorbidities, or scanner conventions. We encourage external validation before downstream use (Section 4.4.4). Additionally, the dataset is imbalanced across histologic subtypes, with clear cell RCC cases substantially outnumbering papillary and chromophobe cases, a distribution that mirrors clinical prevalence but may affect model performance on rarer subtypes. We recommend stratified sampling, class-weighted losses, or augmentation for the rarer chromophobe (n=18) and papillary (n=26) subtypes.

RCC-AID is intended for research purposes only. Models trained on this dataset should not be used for direct clinical decision-making without independent prospective validation on representative local cohorts. We recommend that train/validation/test splits be made publicly available alongside any published results, to facilitate reproducible benchmarking. We do not impose a single reference split given the dataset’s size and the range of possible task definitions (e.g., segmentation and classification). We recommend patient-level splitting to avoid leakage.

Future versions of this dataset may include the extension to MR imaging. We also intend to incorporate any contributed annotations from the research community into versioned dataset updates, with inter-observer variability metrics reported where multiple annotations are available for the same case. The 476 clear cell RCC scans not selected for annotation remain available via TCIA for future annotation efforts, requiring comparable per-case review to Section 5.2.2.

## 4. Resource Availability

### 4.1 Data/Code Location

Images and voxel-level annotations of kidneys and lesions are publicly available without registration at https://zenodo.org/records/20719257 (de Boer et al., 2026). The repository contains one NIfTI file per CT scan, corresponding segmentation masks, and a CSV file with metadata. Code accompanying this dataset can be found on GitHub: https://github.com/DIAGNijmegen/oncology-RCC-AID-codebase.

### 4.2 Potential Use Cases

RCC-AID is suitable for a wide range of medical imaging and machine learning tasks, including:

▪ **Segmentation:** Voxel-level annotations enable training and evaluation of kidney, tumor, and cyst segmentation models.
▪ **Detection:** Voxel-level annotations allow for training and evaluating detection models that detect the primary tumor and or the kidney cysts.
▪ **Classification:** Histologic subtype labels (clear cell, papillary, chromophobe) support subtype classification from CT.
▪ **Radiomics and survival analysis:** Combined with TCGA clinical, genomic, and pathology data, this dataset supports multimodal prognostic modeling.

### 4.3 Licensing

The underlying TCGA imaging data is released under a Creative Commons Attribution 3.0 license (https://creativecommons.org/licenses/by/3.0/). The annotations provided in this work are licensed under a Creative Commons Attribution 4.0 International license (https://creativecommons.org/licenses/by/4.0/), which is compatible with the upstream CC BY 3.0 license. Users may freely use, share, and adapt this dataset for any purpose, including commercial use, provided that appropriate credit is given to both the original TCGA data providers and the authors of this work. The code released alongside this dataset is licensed separately under the MIT license.

#### 4.3.1 Plain-language summary

You can use, modify, and redistribute this dataset freely, including for commercial purposes, as long as you credit the source. You may not claim the data as your own or impose additional restrictions on others who receive it.

### 4.4 Ethical Considerations

#### 4.4.1 Ethics approval

All imaging data used in this study were obtained from the publicly available TCGA repository via TCIA. This data was originally collected under appropriate institutional review board approval and patient consent as part of the TCGA Research Network. No new patient data were collected for this study. The results published here are in whole or in part based upon data generated by the TCGA Research Network: http://cancergenome.nih.gov/.

#### 4.4.2 Informed consent

Participants in the original TCGA study provided broad informed consent permitting secondary research use of their data, including imaging. No direct patient contact or collection of new data was performed in this study.

#### 4.4.3 Data sensitivity and anonymization

The imaging data distributed through TCIA was de-identified by the original data providers per HIPAA Safe Harbor standards, including removal of DICOM identifiers. No additional re-identification steps were applied. We classify this dataset as low-sensitivity.

#### 4.4.4 Intended use and misuse risks

This dataset is intended for non-commercial and commercial research and product development in medical image analysis and machine learning, and should not be used for direct clinical decision-making without regulatory approval and independent validation. Re-identification attempts are prohibited and suspected breaches should be reported to the contact address below.

#### 4.4.5 Official contact

For questions regarding data access, ethical use, or data protection, please contact.

#### 4.4.6 Version policy

If a participant withdrawal request is received or a data quality issue is identified, the affected records will be removed from the repository and a new dataset version will be issued with a corresponding changelog. Users who have downloaded a previous version will be notified via the Zenodo record.

Users of RCC-AID are expected to adhere to applicable research ethics standards, including the prohibition on reidentification, responsible reporting of data issues, and use of the data in accordance with the terms of the license. For guidance, we refer users to the Responsible AI Licenses (RAIL) framework (https://www.licenses.ai/).

## 5. Methods

### 5.1 Data Details

This is a retrospective study based entirely on pre-existing, publicly available imaging data downloaded from TCIA in November 2025. Data from three datasets available from TCGA were used in the dataset creation, TCGA-KIRC (Akin et al., 2016), TCGA-KIRP (Linehan et al., 2016a), and TCGA-KICH (Linehan et al., 2016b). The original collections include multiple image modalities for all patients, plus clinical data, genomics, image analyses, and histopathology information in many cases. The number of patients per dataset and other dataset information are described in Table 1. The data was downloaded from the TCIA using the NBIA Data Retriever (The Cancer Imaging Archive, 2024) on November 14, 2025.

**Table 1:**
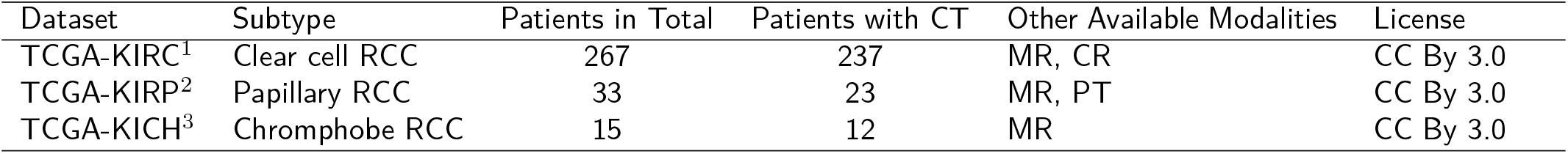
Dataset information for the TCGA RCC cohorts. ^1^ (Akin et al., 2016), ^2^ (Linehan et al., 2016a), ^3^ (Linehan et al., 2016b)

#### 5.1.1 Clinical and molecular variables

In addition to imaging data, the TCGA cohorts provide clinical and molecular information for most patients. Available variables include histologic subtype, tissue slides, overall survival, and genomic profiles. These can be linked to the imaging data via shared TCGA participant identifiers, enabling multimodal analyses. The metadata CSV released alongside this dataset includes a subset of these variables for the 91 patients in the final cohort.

### 5.2 Methods Used for the Data Creation

#### 5.2.1 Data preprocessing and screening

The TCGA kidney collections (KIRC, KIRP, KICH) include multiple CT scans and MRI sequences for each patient. Using an automated script all CT series (n = 1,915) from 272 patients were screened: we filtered for axial scans, *≥*10 slices, slice thickness *≤*5 mm, existing series descriptions, and excluded scout/reconstruction series. Additionally, only the first study acquisition date per patient was retained, to reduce post-surgery scans. In total 759 eligible CT series from 247 patients remained, consisting of mostly clear cell RCC (n = 676), followed by papillary RCC (n = 56), and chromophobe RCC (n = 27). Weighing the substantial annotation effort against the comparatively lower incremental value of additional clear cell RCC cases, we limited clear cell RCC to a random subset of 200 scans, while retaining all available papillary RCC and chromophobe RCC cases to ensure adequate representation of these rarer subtypes. Because subsampling was random rather than stratified, the retained 200 cases should approximate the full distribution, targeted stratified sampling was not performed but could be considered in future annotation efforts. Exclusion criteria are listed in Figure 1.

**Figure 1:**
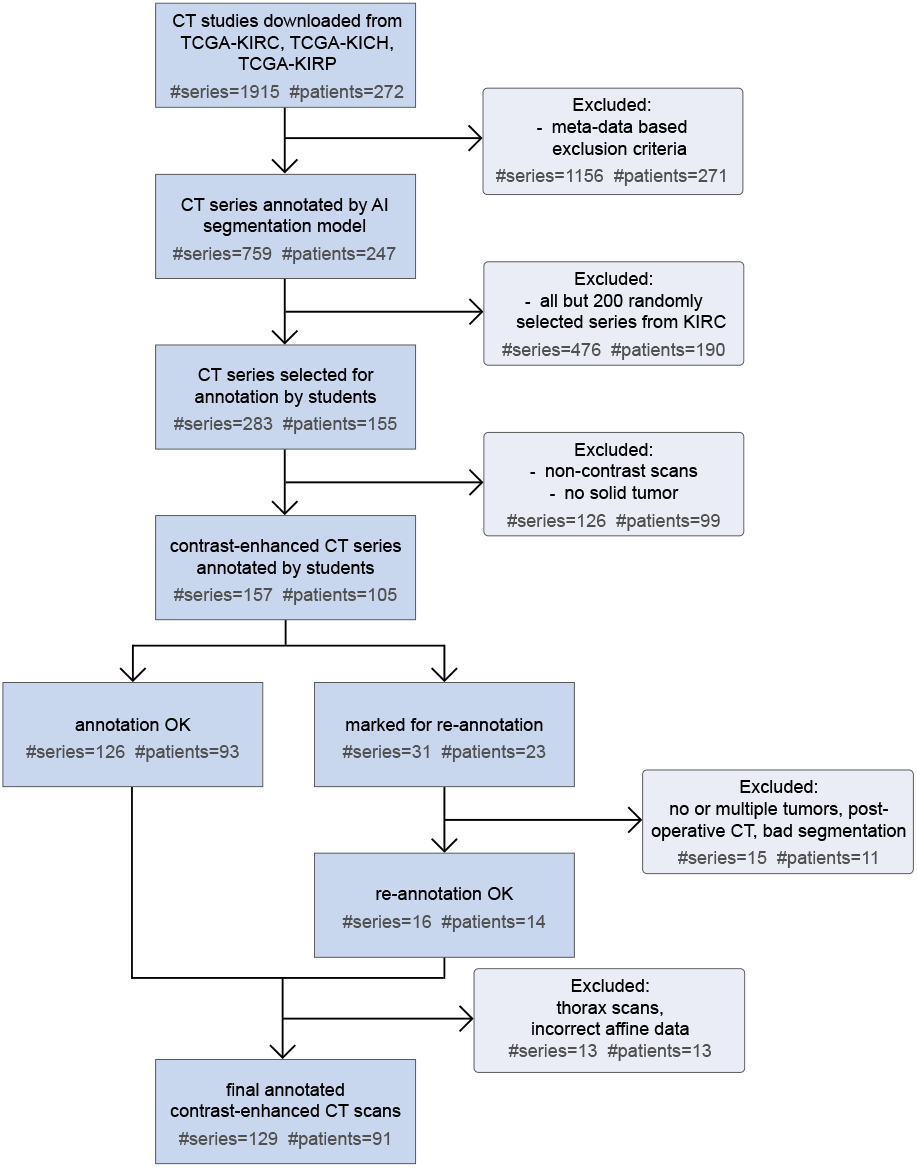
Overview of the RCC-AID inclusion and exclusion process, including the number of series and patients at each stage. Note that patient counts reported in the exclusion boxes refer to the number of patients represented within the excluded data, rather than the number of patients removed from the cohort.

#### 5.2.2 Annotation protocol

The goal of this study was to create a reliable dataset of voxel-wise annotations for solid kidney tumors and cysts. To align with clinical standard and maintain a straightforward annotation pipeline, two inclusion criteria were applied. First, only contrast-enhanced CT was included, as these are routinely used for clinical assessment of renal pathologies. Since TCGA metadata does not provide sufficiently clear or structured information regarding contrast phases, we could not reliably filter for this automatically. Instead, we established a general “contrast-enhanced” requirement and tasked our annotators with manually verifying this during the review process. Second, we restricted RCC-AID to cases containing exactly one solid primary tumor, alongside any complementary cysts. While patients can present with multiple solid lesions, these cases are relatively uncommon and make distinguishing complex cysts from solid tumors highly challenging on CT. We excluded these cases to ensure the final dataset remained consistent and diagnostically certain.

To generate the annotations, we first used a public 3D segmentation model (de Boer et al., 2026) to automatically delineate kidneys and an abstract class “abnormal renal mass” across all selected scans. Next, we set up an annotation pipeline using the reader study functionality of grand-challenge (Meakin et al., 2025). We trained student annotators on the reader study’s functionality and on visual markers distinguishing cysts from tumors. They manually adjusted the lesion segmentation masks if they saw obvious mistakes or missed parts of the lesions. Additionally, they placed click-points on the primary solid tumor and any identifiable cysts. Any unselected lesion masks were subsequently deleted. Unidentified lesions were not checked for or added. Annotators actively flagged and removed any non-contrast scans they encountered. Cases were the annotators were uncertain, or could not identify a primary tumor, were sent for verification to a board-certified radiologist and discarded if no certain selection could be reached. Contrast phases were additionally annotated by a board-certified radiologist.

A total of 283 CT series were included for initial annotation. Of these, 31 cases (11.0%) were referred to a board-certified radiologist for re-annotation due to two reasons. First, 25 cases were directly flagged by students who were uncertain of the tumor subtype. Second, the remaining six cases required an additional review because the recorded click-points could not be successfully mapped to a unique primary tumor. Following this expert review, 16 of the 31 cases were successfully resolved and retained. The remaining 15 cases were excluded from the final dataset. Reasons for exclusion included diagnostic uncertainty due to post-surgical changes (n=6), an unidentifiable tumor (n=5), spatial deviation of click-points and tumor mask (n=2), the presence of multiple tumors (n=1), and substantial errors in the automated segmentation (n=1).

#### 5.2.3 Annotator expertise

Initial quality checks and mask corrections were performed by two students who were trained for the task but did not receive full medical training. Annotators received a standardized briefing on the annotation protocol and the relevant imaging characteristics of RCC subtypes before beginning the review process. Uncertain cases were escalated to a board-certified radiologist with 6 years of experience in abdominal imaging, who made the final inclusion and exclusion decisions. Another board-certified radiologist with 3 years of experience annotated the contrast phases of the CT scans, into either arterial, venous, or delayed.

#### 5.2.4 Inter-observer variability

Formal inter-observer variability was not assessed in this study, as each case was reviewed by a single student annotator with radiologist review for uncertain cases only. However, in a recent study, inter-observer variability on kidney lesions yielded a mean Dice similarity coefficient of 0.664 (±0.274) (Mamani et al., 2023). We encourage researchers who annotate any of the same cases to share their masks, which would enable future inter-observer analyses.

#### 5.2.5 Acquisition equipment and parameters

As TCGA is a multi-site, multi-scanner collection, CT acquisition parameters vary substantially across cases. Scanner manufacturers include SIEMENS (52%), GE MEDICAL SYSTEMS (39%), Philips (6%), and Toshiba (3%). Where available, acquisition parameters such as slice thickness, pixel spacing, and reconstruction kernel are embedded in the DICOM headers of the original data and are summarised in the released metadata CSV. Table 2 summarizes the acquisition variables. All scans are contrast-enhanced, with varying contrast phases. Given the acquisition heterogeneity, we recommend resampling volumes to a common voxel spacing prior to training and reporting performance stratified by acquisition parameters where feasible.

**Table 2:** Acquisition variables by RCC subtype (mean ± standard deviation where applicable). Contrast phases include arterial / venous / delayed.

| Variable | Clear Cell RCC | Papillary RCC | Chromophobe RCC | Total |
| --- | --- | --- | --- | --- |
| N (scans) | 85 | 26 | 18 | 129 |
| Tube voltage (kVp) | 120.4 ± 1.9 | 120.0 ± 0.0 | 120.0 ± 0.0 | 120.2 ± 1.5 |
| Tube current (mA) | 317.3 ± 109.9 | 344.2 ± 126.9 | 303.2 ± 187.3 | 320.8 ± 126.2 |
| Exposure (mAs) | 1147.6 ± 1775.1 | 312.6 ± 821.1 | 183.9 ± 111.4 | 842.5 ± 1540.9 |
| Slice thickness (mm) | 4.0 ± 1.3 | 4.7 ± 0.9 | 3.8 ± 1.5 | 4.1 ± 1.3 |
| Pixel spacing (mm) | 0.8 ± 0.1 | 0.8 ± 0.1 | 0.7 ± 0.0 | 0.8 ± 0.1 |
| Contrast phase | 15 / 35 / 35 | 5 / 9 / 12 | 6 / 6 / 6 | 26 / 50 / 53 |

#### 5.2.6 Data format

All images and segmentation masks are provided in NIfTI format (.nii.gz). Each case consists of one CT volume and one corresponding multi-label segmentation mask, where label values encode background (0), kidney (1), solid tumor (2), and cyst (3). Patient-level metadata is provided as a structured CSV file. A README file in the repository describes the complete file naming convention and folder structure.

## 6. Validation

### 6.1 Dataset summary

RCC-AID includes 129 CT scans (ccRCC n=85, papillary RCC n=26, chromophobe RCC n=18) from 91 patients (24 female, 67 male, mean age 56 years, min. 26, max. 82, one patient’s age is unknown). All CTs are contrast-enhanced with 26 arterial, 50 venous, and 53 delayed-phase scans. The average tumor size is 167 ml, with an average axial diameter of 54 mm. Across 38 out of the total 129 CTs, there are 118 cysts, the average cyst size is 17 ml, with an average diameter of 21 mm. The tumor characteristics per subtype are described in Table 3. Figure 2 presents per subtype the smallest and largest tumor volume, highlighting the difficulty of the dataset with extremely large but also tiny tumors. In Figures 3a and 3b the distributions of tumor volume and diameter per subtype is presented, consistent with literature: 70% of patients present with stage I (diameter *≤*7cm) and 11% with stage IV RCC (Rose and Kim, 2024). Figure 3c shows a low correlation between tumor volume and age (Pearson’s coefficient r=0.03). Figure 3d shows a strong correlation between tumor volume and diameter (presented in log scale with Spearman’s coefficient r=0.99). Lastly, Figures 3e and 3f present the age distribution for male and female patients and the distribution of sex per subtype respectively, with the literature reporting male-to-female ratio of 1.5:1.0 and incidence peak at age 60-70 years (Capitanio and Montorsi, 2016).

**Table 3:**
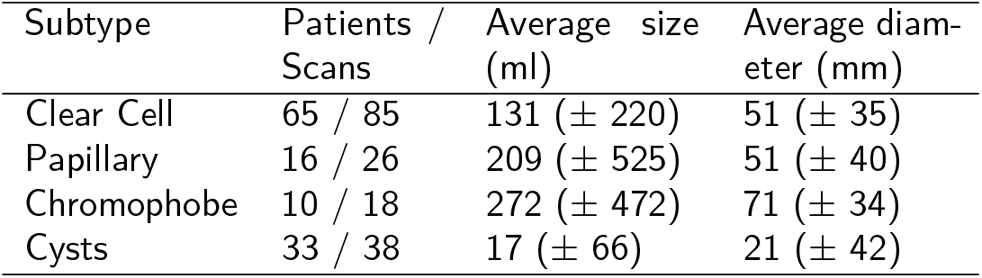
Macro-average tumor size and diameter per RCC subtype/cyst, the cysts are present alongside the RCC sub-types and are not additional cases. Standard deviation is presented in parentheses.

**Figure 2:**
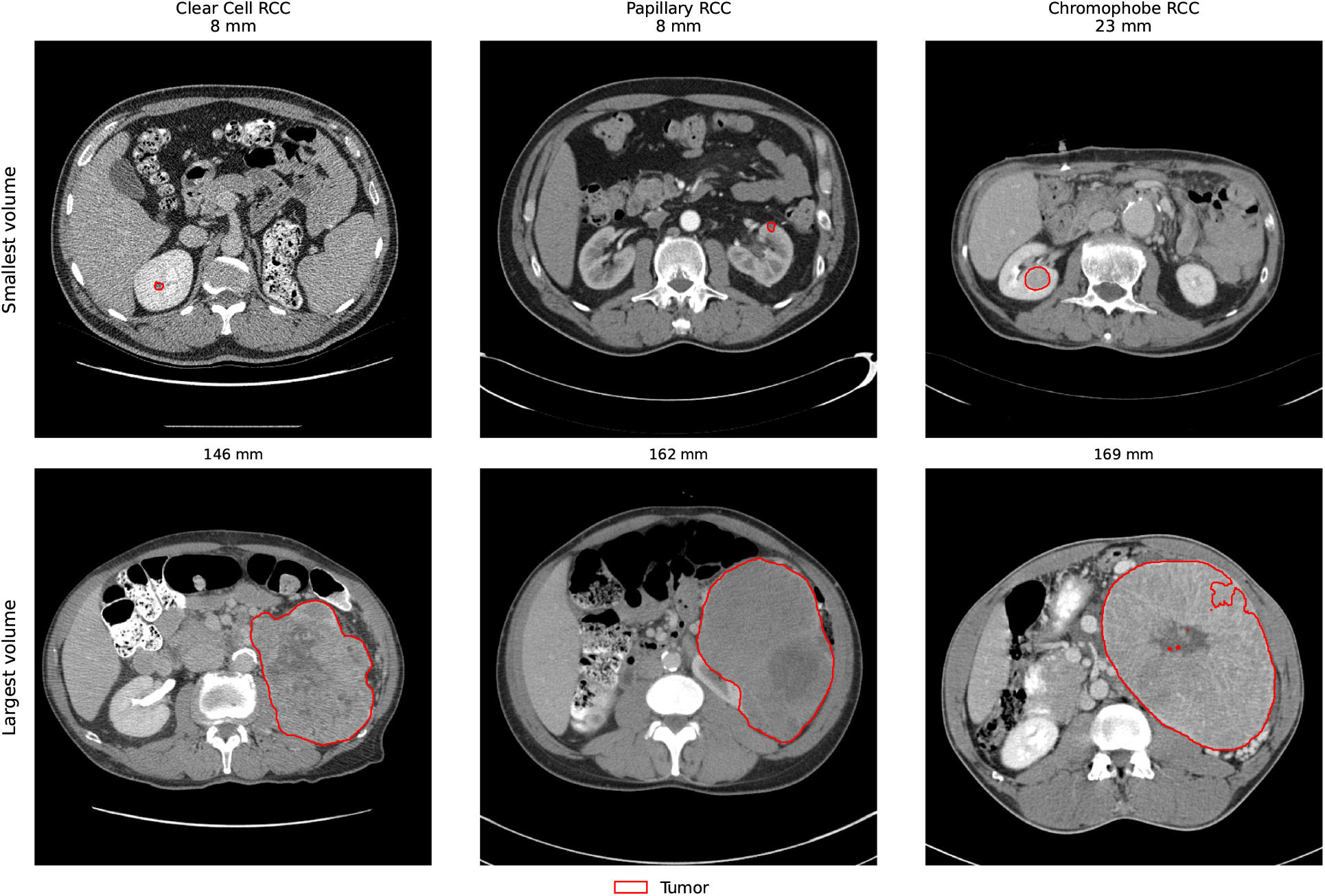
Extreme cases in the RCC-AID: smallest and largest tumors for the three different pathologies. Maximum tumor diameters are noted in the subfigure titles.

**Figure 3:**
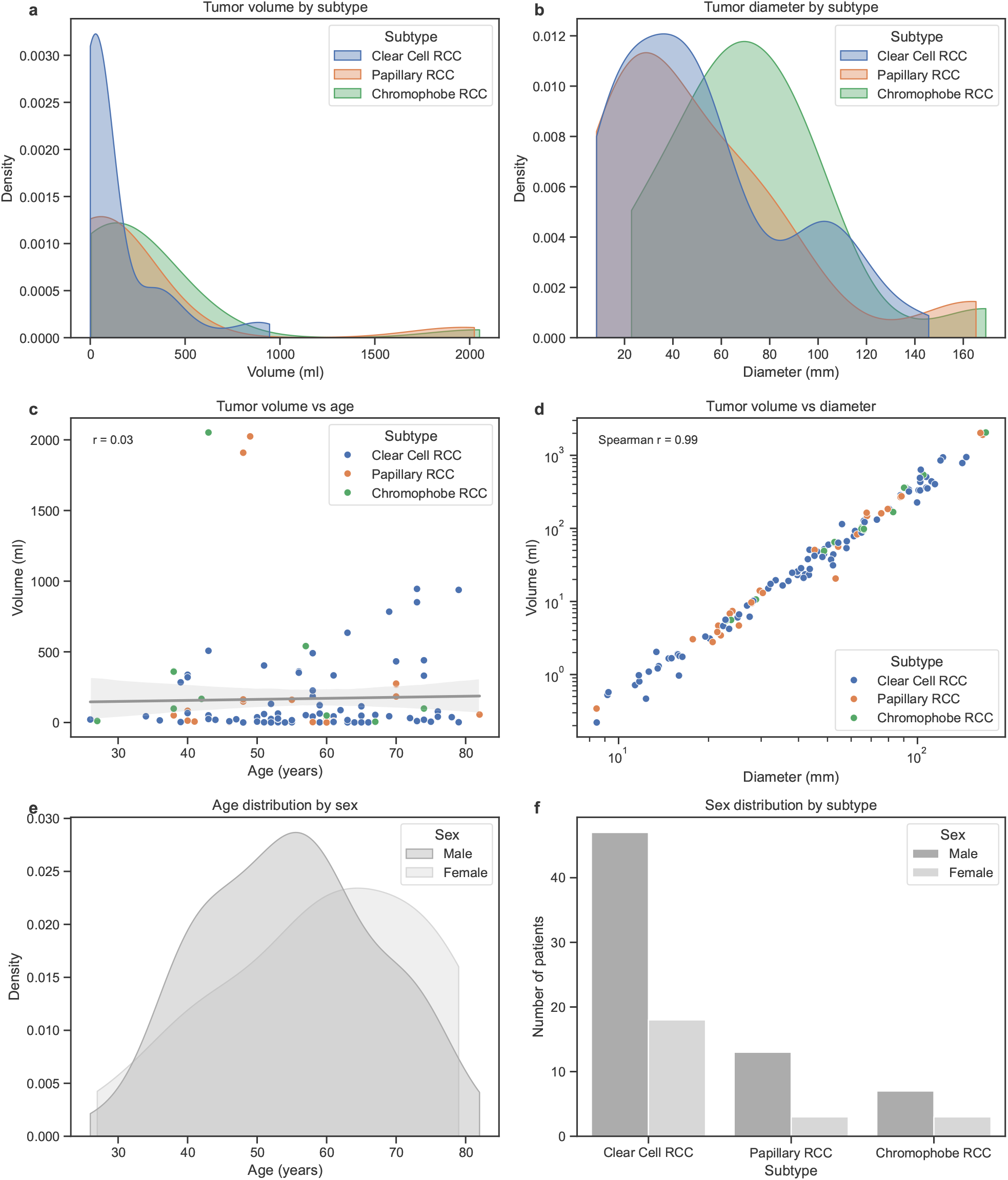
Dataset analysis. **a** and **b** present the tumor volume (in ml) and tumor diameter (in mm) distributions per tumor histologic subtype as Kernel Density Estimation (KDE) plots. **c** presents the correlation between tumor volume (in ml) and patient age, the Pearson correlation coefficient (r) is presented in the plot. **d** presents the correlation between tumor volume (in ml) and tumor diameter (in mm) with log scale, the Spearman correlation coefficient (r) is presented in the plot. **e** presents the age distribution for male and female patients as a KDE plot. Finally, **f** presents the sex distribution per tumor histologic subtype.

### 6.2 Annotation reliability and quality control

A final manual quality assurance check was performed on all volumes remaining after exclusion criteria had been applied. Six thoracic scans were excluded to strictly limit the dataset’s anatomical scope to the abdominal region. Additionally, seven cases were removed due to affine matrix corruptions that caused spatial mismatches between images and annotations.

### 6.3 Example use case

RCC-AID was used in Häntze et al. (2026) for external validation of a renal lesion classification task. The study aimed to distinguish renal masses on CT imaging. Using the RCC-AID segmentations for lesion localization, automatic extraction of individual lesions was performed prior to classification. A total of six model configurations were evaluated, including four foundation-model-based methods, a ResNet trained from scratch, and a radiomics-based classifier. Among these, the radiomics model achieved the best performance, reaching AUC values of 0.93 for cysts, 0.84 for clear cell RCC, 0.90 for papillary RCC, and 0.86 for chromophobe RCC. In comparison, the foundation-model-based approaches showed substantially lower performance, highlighting the challenges of transferring general-purpose pretrained models to a specialized renal lesion task.

## Data Availability

All data produced are available online at https://zenodo.org/records/20719257.

## 8. Acknowledgements

This research is funded by the European Union under HORIZON-HLTH-2022: COMFORT (101079894). Views and opinions expressed are however those of the author(s) only and do not necessarily reflect those of the European Union or European Health and Digital Executive Agency (HADEA). Neither the European Union nor the granting authority can be held responsible for them. The results published here are in whole or part based upon data generated by the TCGA Research Network: http://cancergenome.nih.gov/.

